# Organophosphate and organohalogen flame-retardant exposure and thyroid hormone disruption in a cohort of female firefighters and office workers from San Francisco

**DOI:** 10.1101/2020.10.06.20207498

**Authors:** Jessica Trowbridge, Roy Gerona, Michael McMaster, Katherine Ona, Cassidy Clarity, Vincent Bessonneau, Ruthann Rudel, Heather Buren, Rachel Morello-Frosch

## Abstract

**Background:** Occupational exposures to flame retardants (FR), which are suspected endocrine disrupting compounds, may be of particular concern for firefighters as they are commonly found in consumer products and have been detected in fire station dust and firefighter gear.

**Objectives:** The Women Workers Biomonitoring Collaborative is a community based participatory research study that sought to measure environmental chemicals relevant to firefighting and evaluate their effects on thyroid hormone levels.

**Methods:** We measured 10 FR or their metabolites in urine of female firefighters and office workers from San Francisco: bis(1,3-dichloro-2-propyl) phosphate (BDCPP), bis(2-chloroethyl) phosphate (BCEP), dibutyl phosphate (DBuP), dibenzyl phosphate (DBzP), di-p-cresyl phosphate (DpCP), di-o-cresyl phosphate (DoCP), 2,3,4,5-tetrabromobenzoic acid (TBBA), tetrabromobisphenol-A (TBBPA), 5-OH-BDE 47, and 5-OH-BDE 100. We assessed potential predictors of exposure levels and the association between FR exposures and thyroxine (T_4_) and thyroid stimulating hormone (TSH).

**Results:** BDCPP, BCEP, and DBuP were the most commonly detected FRs, among all study participants, with intermediate BMI and college educated women having the highest levels, and Black women having higher BDCPP levels than White women. Firefighters had higher detection frequencies (DF) and exposure levels compared to office workers; median BDCPP levels were five times higher in firefighters than in office workers. Among firefighters, occupational activities were not significantly associated with FR levels, although position (i.e. officer and firefighter versus driver), being on-duty (versus off-duty) and assigned to the airport suggested a positive association with FR levels. Among firefighters, a doubling of BDCPP was associated with a 2.88% decrease (95%CI −5.28,-0.42) in T_4_. We did not observe significant associations between FR and T_4_ among office workers.

**Discussion:** Firefighters had significantly higher exposures to FR compared to office workers, and we observed a negative association between BDCPP and thyroxine in firefighters. Future research should elucidate occupational sources of FR exposure and opportunities for exposure reduction.

## Introduction

Studies show that firefighters have elevated risk for many cancers. For instance, a meta-analysis of 32 studies identified increased rates of lymphoma, and testicular and prostate cancer among male firefighters (LeMasters et al. 2006). A study of over 19,000 male firefighters in a pooled cohort from San Francisco, Chicago and Philadelphia found that the occupation of firefighting was associated with elevated lung cancer incidence and mortality, and leukemia mortality (Daniels et al. 2015). Breast cancer incidence was non-significantly elevated among men and among the 991 women included in the study and in both groups the increases were largest among those at younger ages (<65 years old for men and ages 50-55 for women (Daniels et al. 2014)). In a published update adding 7 years of follow up to the pooled cohort, Pinkerton et al. (2020) found that women firefighters had elevated mortality rates for bladder cancer and non-statistically elevated mortality from Non-Hodgkin’s lymphoma, multiple myeloma, lung cancer and breast cancer (Pinkerton et al. 2020). A study of Florida firefighters that included 5,000 women found elevated incidence of cervical and thyroid cancer and Hodgkin’s disease compared to the general Florida population (Ma et al. 2006) and a more recent study of Florida firefighters using that state’s cancer registry found that women firefighters had elevated risk of melanoma, thyroid, and brain cancer (Lee et al. 2020).

In addition to limited studies of cancer risk among women firefighters, few studies have assessed their exposures to occupational hazards, including environmental chemicals, although occupational exposures have been well documented among male firefighters. Common exposures measured in firefighters and fire stations include polyaromatic hydrocarbons (PAH), formaldehyde, benzene, dioxins, diesel, per- and poly-fluoroalkyl substances (PFAS), and flame retardants including organohalogen flame retardants, like polybrominated diphenyl ethers (PBDE), and organophosphate flame retardants (OPFR) (Caux et al. 2002; Dobraca et al. 2015; Fent et al. 2014; Grashow et al. 2020; Jin et al. 2011; Laitinen et al. 2014; Park et al. 2015; Shaw et al. 2013; Shen et al. 2015, 2018; Trowbridge et al. 2020). Many of these chemicals have demonstrated their potential for breast tumor development in animal and human studies (Rodgers et al. 2018; Rudel et al. 2011, 2014) and their capacity as endocrine disrupting compounds (EDC) (Gore et al. 2015; Rudel et al. 2011).

Flame retardants are of particular interest for firefighters because they have been found in firefighting gear (Alexander and Baxter 2016) and in fire station dust (Shen et al. 2015, 2018). Additionally, firefighter biomonitoring and studies of fire station dust samples have found elevated levels of brominated flame retardants compared to homes and offices (Park et al. 2015; Shen et al. 2015). One study found that dust collected from 26 fire stations across the US had elevated levels of tris(1,3-dichloro-isopropyl)-phosphate (TDCPP), an OPFR flame retardant, at levels higher than those measured in homes and offices (Shen et al. 2018).

California’s furniture flammability standard known as TB117 was in effect between 1977 and 2013 and required furniture filling such as foam to withstand an open flame test without igniting. In order to meet the standard, manufacturers typically added chemical flame retardants to filling materials such as flexible polyurethane foam (PUF). Additionally, furniture manufacturers often complied with the California standard for their products sold nationwide, impacting everyone in the U.S. (Castorina et al. 2017; Dodson et al. 2014; Stapleton et al. 2012). Because flame retardants are added to products post-production and are not bound to fabric and foams, they can leach out of materials and contaminate air and dust (Dodson et al. 2012; Stapleton et al. 2012). California’s revised furniture flammability standard TB117-2013 removed the open flame test in favor of a smolder test designed to mimic a lit cigarette on furniture fabric, and manufacturers have been able to meet this standard without adding chemical flame retardants in PUF (Bureau of Electronic and Appliance 2018). At the same time, concerns about toxicity and bioaccumulation of PBDE flame retardants in the early 2000s led to an increase in the use of other flame retardants such as OPFRs in both furniture and fabrics (van der Veen and de Boer 2012a). Although use of flame retardants in furniture has declined, many are persistent and bioaccumulative, and both legacy and replacement FR remain in durable consumer products, such as furniture, and exposures remain a concern (Dodson et al. 2012; Zota et al. 2013). OPFRs are commonly used as flame retardants in fabric, electronics, PUF, as plasticizers, and engine lubricants (Covaci et al. 2011; Van den Eede et al. 2012; van der Veen and de Boer 2012a). Due to OPFR use in fabrics and that PBDEs have been found in firefighter gear (Alexander and Baxter 2016), OPFRs may conceivably be used in firefighter clothing and protective gear. Organophosphate and brominated flame retardants may be associated with cancer (Lerro et al. 2015; Rudel et al. 2014) and have also been identified as endocrine disruptors that are associated with altered thyroid hormone (TH) levels in both *in vitro* and *in vivo* studies (**Table 1**) (Dishaw et al. 2014; Farhat et al. 2013; Hill et al. 2018; Meeker and Stapleton 2010; Wang et al. 2013b).

**Table 1.**
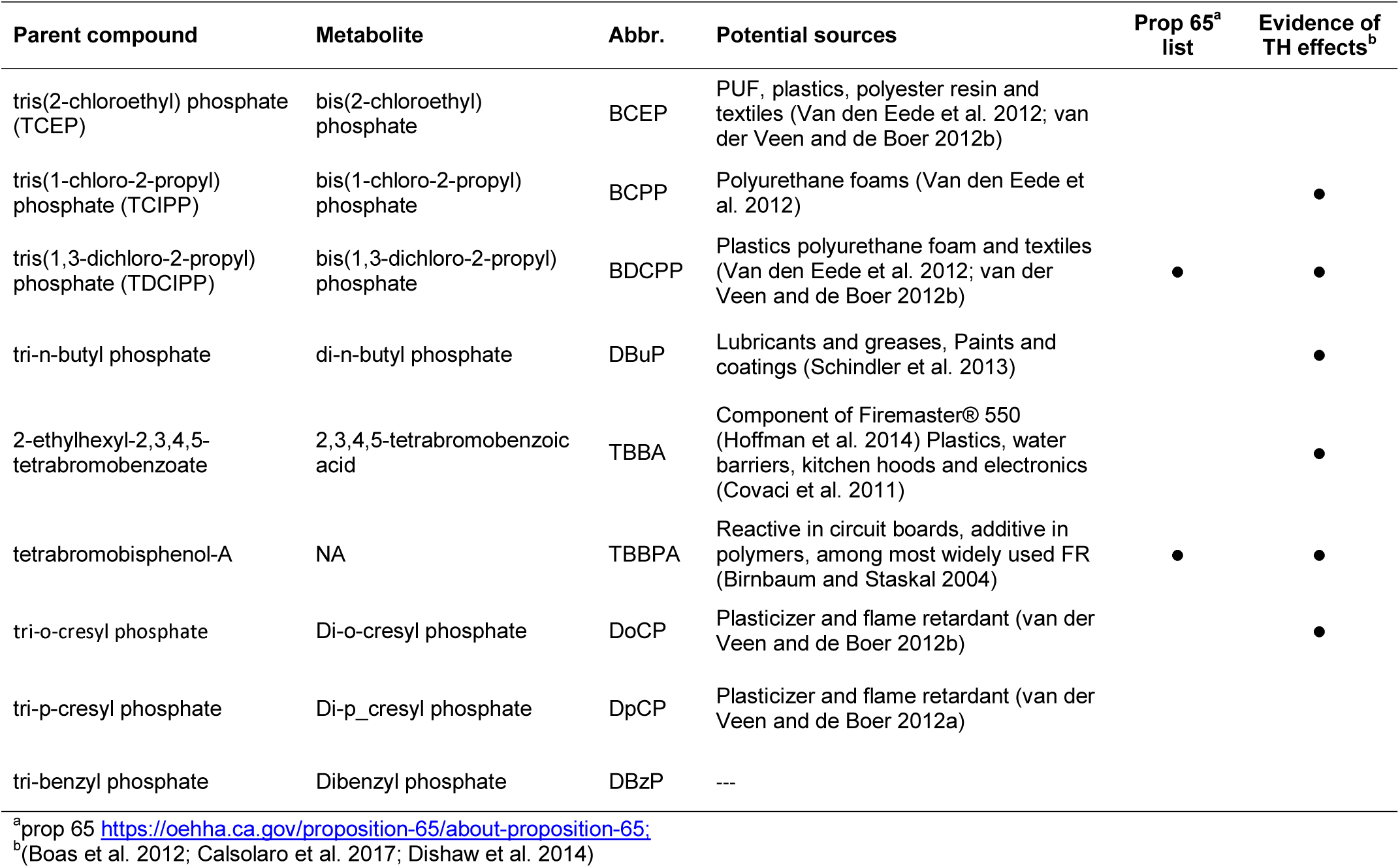
Select flame retardant compounds, their metabolites, potential sources and uses and evidence of toxicity

### Thyroid hormone disruption

Studies suggest that TH levels, including thyroid stimulating hormone (TSH) and thyroxine (T_4_), are affected by exposure to environmental chemicals including OPFRs (Diamanti-Kandarakis et al. 2009; Dishaw et al. 2014; Gore et al. 2015; Rudel et al. 2014). TH production is controlled via a negative feedback loop: as TH levels decline in the body, the pituitary gland secretes TSH, which induces an increase in production of T_4_ by the thyroid gland. TSH secretion is suppressed when T_4_ levels reach a “set point” that varies by individual (Zoeller et al. 2007).

Studies of zebrafish and chicken embryos show that OPFRs can disrupt thyroid hormone homeostasis and decrease total T_4_ concentrations (Farhat et al. 2013; Wang et al. 2013a). Human studies suggest impacts of exposure to OPFRs on thyroid hormone function, although research remains limited and the direction of the association is inconclusive. Meeker and Stapleton found decreased T_4_ levels with higher TDCPP concentrations in house dust among men (Meeker and Stapleton 2010). A study of e-waste recycling workers found that a two-fold increase in exposure to tert-butyl diphenyl phosphate was associated with lower total T_4_ levels in men (Gravel et al. 2020); and Preston et al. (2017) found that high levels of diphenyl phosphate, a metabolite of triphenyl phosphate, were associated with an increase in total T_4_ concentration in women (Preston et al. 2017). Thyroid hormones are important for fetal neurodevelopment and for TH-mediated gene expression. Additionally, TH may be relevant to downstream adverse health impacts such as cardiovascular disease and cancer (Gore et al. 2015; Krashin et al. 2019; Rodondi et al. 2006).

While biomonitoring studies show that firefighters have higher body burdens of flame retardant chemicals than the general United States population (Alexander and Baxter 2016; Brown et al. 2014; Park et al. 2015), very little research has investigated the extent and health implications of exposure among women firefighters due to the limited number of women in most fire departments. Assessing the potential health risks of flame-retardant exposures among women firefighters, including for outcomes such as breast cancer poses methodological challenges due to the low numbers of women in the fire service and the long latency period from exposure to onset of disease. Assessing thyroid hormone disruption associated with flame retardant exposures enables identification of early biological perturbations of potential relevance to thyroid dysfunction or disease (Ward et al. 2010), and long-term adverse health outcomes such as cancer (Krashin et al. 2019).

San Francisco has one of the largest forces of women firefighters among large urban fire departments in the U.S.—approximately 15% of firefighters in SFFD are women. As firefighting and other first responder professions continue to diversify and increase the number of women in their ranks, it is important to understand occupational exposures and potential health implications for women firefighters. The Women Workers Biomonitoring Collaborative (WWBC) is a community-based participatory research study that aims to characterize occupational exposure to potential breast carcinogens among women workers. This study sought to characterize exposures to OPFRs and replacement organohalogen flame retardants (henceforth referred to together as flame retardants (FR)) among women firefighters and office workers in San Francisco and to assess exposure to FR and their effect on total T_4_ and TSH.

## Methods

Study design and participant recruitment protocols have been described elsewhere (Grashow et al. 2020; Trowbridge et al. 2020). Briefly, participant recruitment, interviews and sample collections took place between 2014 and 2015. Participants were employees of the City and County of San Francisco or the San Francisco Fire Department (SFFD). Firefighter collaborators and researchers actively recruited study participants through the Fire Department, as well as firefighter advocacy organizations including the San Francisco Firefighters Cancer Prevention Foundation, United Fire Service Women, and the International Association of Firefighters Union Local 798. Office workers, who are non-first responder employees of the City and County of San Francisco, were recruited by emails to employees city-wide, tabling, and presentations by research staff and firefighter collaborators. Study participants were eligible to participate if they were 18 years or older and non-smokers. Firefighters were required to have worked in SFFD for a minimum of 5 years and be on “active duty” (i.e. currently assigned to a fire station at the time of recruitment). Participants were consented into the study following protocols approved by the Institutional Review Board of the University of California, Berkeley (# 2013-07-5512).

### Data collection

All participants completed an hour-long in-person exposure assessment interview with research staff. This interview collected demographic and basic health information including body mass index (BMI) and the use of hormone replacement medications. We also asked about possible sources of FR exposure including consumer product use, diet, and occupational activities. A subset of participants (N = 66) gave researchers permission to access their departmental firefighting history records, from which we abstracted the number of fires fought in the 7 days and month prior to the sample collection date.

We collected most biospecimen samples, including blood and urine, between 8AM and 11AM. A trained phlebotomist collected blood in EDTA-treated lavender top tubes. Urine specimens were collected by participants in 60mL polypropylene biospecimen cups. Prior to sample collection, we gave participants a biospecimen cup and instructions for collecting a morning void sample (first urine sample after a night’s sleep) which they brought with them to their sample collection appointment. Research staff then asked for a second, spot urine, sample at the time of the blood collection. Biospecimens were put on ice and transferred to the lab at the University of California, San Francisco where research staff processed samples within 3 hours of their collection. Blood collection tubes were spun at 3000 rpm for 10 minutes and plasma was aliquoted into 1.1 mL cryovial tubes. Urine samples were aliquoted into 3.5 mL cryovial tubes. All samples were stored at −80°C until analysis.

### Laboratory analysis

#### FR analysis

We sought to quantify metabolites in urine of six OPFR chemicals: bis(1,3-dichloro-2-propyl) phosphate (BDCPP), bis(2-chloroethyl) phosphate (BCEP), dibutyl phosphate (DbuP), dibenzyl phosphate (DBzP), di-p-cresyl phosphate (DpCP), di-o-cresyl phosphate (DoCP), and 4 brominated flame retardants: 2,3,4,5-tetrabromobenzoic acid (TBBA), tetrabromobisphenol a (TBBPA), 5-OH-BDE 47, and 5-OH-BDE 100.

Quantification of the 10 analytes was performed using liquid-chromatography-tandem mass spectrometry (LC-MS/MS) on an Agilent LC 1260 (Agilent Technologies, Sta. Clara, CA)-AB Sciex 5500 system (Sciex, Redwood City, CA). Freshly thawed urine specimens (1 mL) were deconjugated prior to LC-MS/MS analysis by addition of 450 U *H. pomatia* glucuronidase (Sigma-Aldrich, St Louis, MO) and incubated at 37 °C for two hours with constant shaking. Deconjugated urine samples were then prepared for LC-MS/MS analysis by solid phase extraction (SPE) using Waters Oasis WAX cartridges (10 mg, 30 μm, 1 cc). An Agilent ZORBAX Eclipse XDB-C8 column (2.1×100 mm, 3.5um) maintained at 50°C was used in reversed-phase chromatography. The analytes were separated by gradient elution using water with 20 mM ammonium acetate as mobile phase A (MPA) and acetonitrile as mobile phase B (MPB). The gradient used for analyte separation consisted of 5% MPB at 0–0.5 min, gradient to 75% MPB from 0.5 to 7.5 min, gradient to 100% MBP from 7.5-9 min, 100% MPB at 9-11 min, and 5% MPB at 11.1–15 min. The analytes were ionized in the negative mode using electrospray ionization (ESI) and mass scanning was performed by multiple reaction monitoring. Each analyte was monitored using two transitions and retention time. Quantitation of each analyte was performed by isotope dilution method with their deuterated or C-13 isotopologues as internal standards.

Each batch of samples was injected in duplicate. Procedural quality control materials and procedural blanks were run along with the calibration curve at the start, middle, and end of each run. Two QC materials were used at low and high concentrations. To accept the results of a batch run, QC materials measurements must be within 20% of their target values and the precision of their measurements have ≤20% CV (coefficient of variation). Analyte identification from total ion chromatograms was evaluated using AB Sciex Analyst v2.1 software while quantification of each analyte was processed using AB Sciex MultiQuant v2.02 software. Analysts were blinded to firefighter and office worker status of the urine samples during the analysis.

#### T_4_ and TSH measurement

The TSH and T_4_ levels were measured in blood plasma using ELISA (Antibodies-online, cat. No. ABIN2773773) following manufacturer’s protocol (Antibodies-online 2020). Room temperature calibrators, controls and samples (25 µl for T_4_ and 50µl for TSH) were loaded onto streptavidin coated wells followed by the addition of biotinylated antibody for T_4_ or TSH. Standard curves were constructed in duplicate with calibrators supplied in the kit. The reaction was incubated at room temperature for 1 hr. and then washed three times. Substrate solution was added and stopped after 15 minutes. Absorbance at 450 nm was read immediately using a microplate reader. The T_4_ and TSH concentrations of each sample, run in duplicate, were obtained from the standard curve. According to the manufacturer’s instructions, samples below the LOD were rerun with a 30 min development time.

### Statistical analysis

The goal of this analysis was twofold: First to characterize exposure to FR chemicals and identify potential predictors of exposure among firefighters and office workers. Second, to assess the relationship between FR exposures and T_4_ and TSH.

We tested the distributions for FRs, T_4_ and TSH visually inspecting the distributions and with the Shapiro-Wilks test. Because of the evidence for skewed distributions, we used nonparametric approaches to test differences between groups (permutation, Wilcoxon) and we natural log-transformed values which improved normality for use in linear models. All regression models were adjusted for log-transformed creatinine to account for urine dilution when quantifying the FR (Barr et al. 2005). We used two different linear regression models based on whether FR levels were being evaluated as the outcome or exposure and to account for FR levels below the limit of detection (LOD). In bivariate analyses looking at the relationship between covariates (e.g. food consumption or position in the fire department) and FR levels as the outcome, we used the maximum likelihood estimation (MLE) model from the NADA package in R, which accounts for levels below the LOD without the need for substitution, when the chemical measurement is the outcome (Helsel 2005). To assessed the relationship between FR levels (exposure) and thyroid hormone (outcome), we used ordinary least squares (OLS) regression models and operationalized levels below the LOD in the following ways: We included all LC-MS/MS reported values (even if those values reported were below the LOD) and substituted 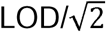 for any remaining non-detect values.

First, we treated FR as the outcome to assess differences between firefighters and office workers and evaluate predictors of FR levels. We calculated summary statistics for FRs including the geometric mean (GM), Geometric Standard Deviation (GSD), and distribution percentiles for each group. We plotted FR levels from WFBC firefighters and office workers along with FR levels from women ages 18-65 from the National Health and Nutrition Examination Survey (NHANES) (2013-2014) to see how WFBC participants compared with a nationally representative sample of the U.S. population. In the full cohort we assessed the impact of variables collected from the exposure assessment interview on FR levels as the outcome using MLE regression. We limited analyses to FRs with a detection frequency (DF) of 70% or higher in at least one group (firefighters or office workers) and applied separate MLE regression models on each FR chemical (continuous outcome) controlling for occupation and log(creatinine). We analyzed the relationship of FR concentrations and the following variables: age, race/ethnicity, body mass index (BMI), and educational attainment and we assessed the association of eating certain foods and packaged foods based on prior literature suggesting an association with FR exposures (Kim et al. 2020).

We then limited the analysis to firefighters to explore specific occupational activities that might be associated with FR levels such as the participant’s assigned position in the fire department (i.e. firefighter, officer, or driver), the frequency of using a self-contained breathing apparatus (SCBA) during fire suppression, salvage and overhaul, the frequency of showering or washing up after a fire event, and the number of fires fought in the week and month prior to the sample collection. We applied MLE regression and exponentiated the beta coefficients and 95% CI to find the proportional change in geometric mean for each unit increase or category change versus referent.

Next, we assessed the impact of FR (exposure) on TH (outcome) using OLS regression and substitution for FR values below the LOD. We ran separate models for each FR chemical and each TH outcome, TSH and T_4_. We considered variables to adjust for in our models if they demonstrated a statistically significant association (p-value < 0.05) with the exposure (at least one FR) and the outcome (TSH or T_4_) in our data or if previous literature identified an association. Although age was not associated with FR in our data, it is associated with TH levels in other studies (Hollowell et al. 2002) and therefore it was included as a covariate in regression models. We did not control for body-mass index (BMI) in regression models because literature suggests that both FRs and thyroid hormone disruption can lead to increased BMI (Boyle et al. 2019; Knudsen et al. 2005), implying that it may be a collider for which adjustment could induce a spurious association. Final models were adjusted for age and log(creatinine). Due to large differences in DF between firefighters and office workers, we stratified the analysis by occupation. We used the continuous exposure when the DF 70% and we categorized FR values when DF < 70%. FR was categorized into the following groups: <LOD and ≥LOD (for FR with DF of 25% to 50%) and <LOD, LOD to median, and >median (for FR with DF between 50% up to 70%). Compounds with DF below 25% were excluded from the multivariate analysis.

Results from the OLS regression models with continuous exposure and outcome were converted to the percent change in the outcome for a twofold increase in FR exposure with the formula: (2^*β*^ - 1) * 100. From OLS regression models with categorical FR exposure and continuous outcomes we calculated the percent change of the outcome for each category compared to the referent (< LOD) with the formula: (e^*β*^ - 1) * 100.

Analyses were conducted using R version 3.6.1 and R-studio version 1.2.1335 (R Core Team 2015; RStudio Team 2016).

## Results

We had chemical measurements for 170 participants and TH measurements for 168 participants. This difference in sample size was due to limited biospecimen samples available to measure TH. We retained the full cohort when characterizing FR levels among firefighters and office workers and assessing the relationship between FR and covariates. When we assessed the relationship between FR levels and TH, we excluded the participants without TH measurements and three participants who reported taking TH replacement medications. The final number analyzed for thyroid hormone disruption was N = 165 (84 firefighters and 81 office workers).

### Flame retardant levels and predictors of exposure

BDCPP had the highest DF and was found in all firefighters (100%) and almost all office workers (90%). BCEP and DBuP were detected in most firefighters (DF>70%) while among office workers the DF was below 50%. TBBPA was detected in 45% of firefighters and 42% percent of office workers while DpCP was detected in 40% of firefighters and 17% of office workers. TBBA, DoCP and DBzP was found in fewer than 30% of firefighters and office workers. OH-BDE and OH-BDE47 were not detected above the LOD in any of our participants’ urine samples. We plotted the distribution (median, interquartile range (IQR) and 95^th^) of each FR detected in our cohort (**Figure 1**; GM (GSD) and percentiles listed in **Table S1**). Firefighters had both higher detection frequencies and higher average levels of FR chemicals compared to office workers with the largest median differences observed for DBuP, BDCPP and BCEP compared to office workers. Indeed, BDCPP levels in firefighters were five times higher than the levels measured in office workers. When compared to NHANES, firefighters had higher levels of DBuP, BDCPP, and BCEP, while office workers had levels similar levels of BDCPP, but lower levels of DBuP and BCEP than NHANES women (**Figure S1**).

**Figure 1:**
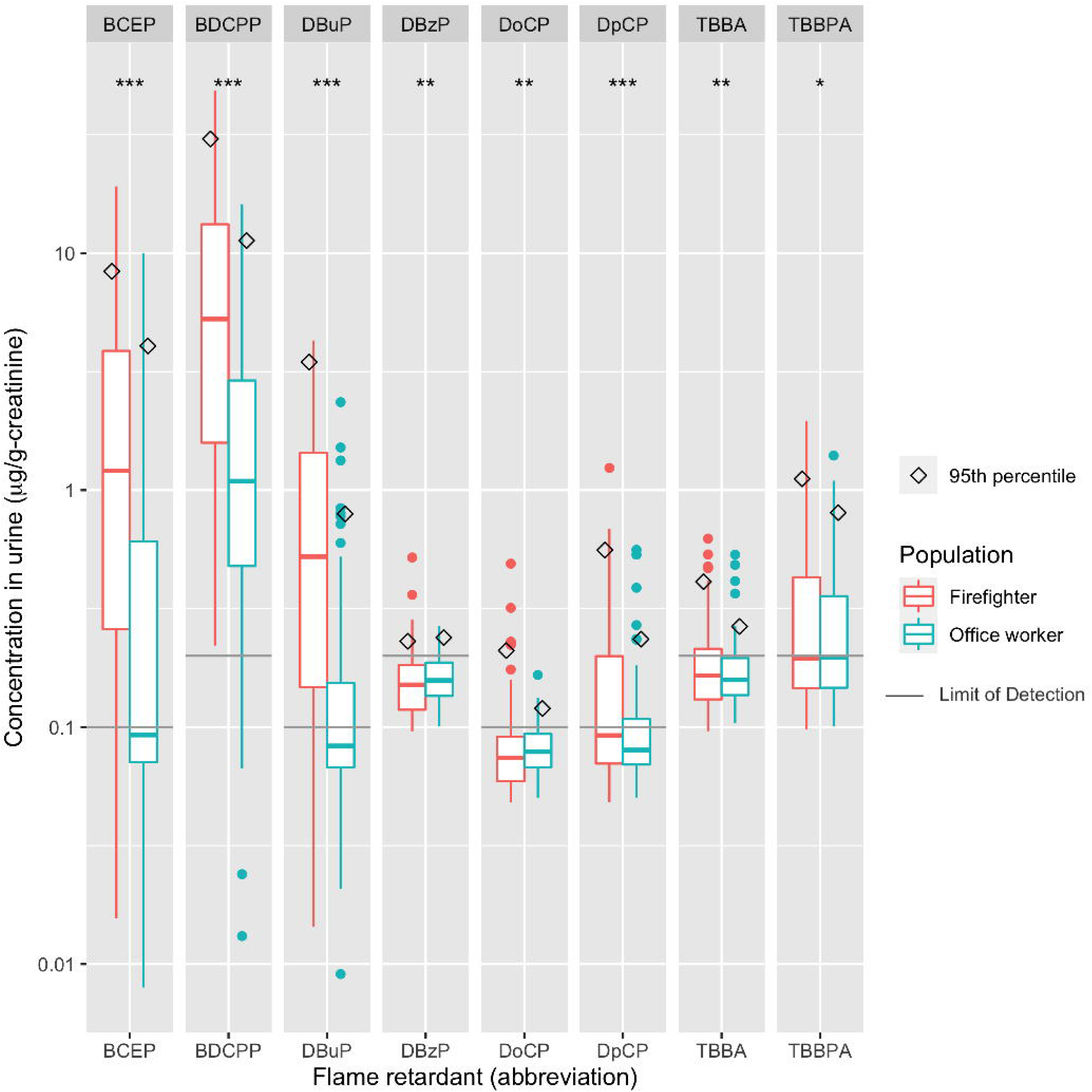
Distribution (median, interquartile range, and 95th percentile) of flame-retardant metabolite levels (µg/g-creatinine) in urine from 86 firefighters and 84 office workers of the WFBC (2014-15). We substituted values below the limit of detection (LOD) with 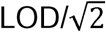. Significance stars represent the p-value from permutation test of the difference in average chemical level between firefighter and office workers: *** <0.001; ** <0.05; *<0.1.

We used MLE regression models to assess the relationship between demographic variables and FR levels, controlling for occupation and log(creatinine) (**Table 2**)These models focused on BDCPP, BCEP and DBuP since these compounds had sufficient detection frequency (DF>70% in at least one group) to use in regression models. We found that having a BMI of 25.0 to 29.0 as compared to 18.5 to 24.9 was associated with increased levels of BDCPP, BCEP, and DBuP (exp(*β*) (95%CI) = 1.71 (1.02, 2.84), 2.70 (1.06, 6.84) and 2.45 (1.28, 4.86) respectively). Likewise, higher education was associated with FR levels; those who completed a bachelor’s degree or higher education had higher BDCPP (exp(*β*) (95%CI) = 1.65 (1.00, 2.73); BCEP, 3.63 (1.49, 8.84); and DBuP, 2.24 (1.17, 4.31)) compared to participants who had completed some college or less. Race and ethnicity were not associated with most OPFRs except for BDCPP, where Black participants had a 2.52 (95%CI 1.10, 5.75) times higher geometric mean compared to White participants.

**Table 2.** Description of covariates among firefighters and office workers and the adjusted ^a^ proportional change in geometric mean (95% CI) of urinary chemical concentration (ng/mL) (DF>70%) for each unit increase, or each category increase compared to the referent, from maximum likelihood estimation (MLE) models

| Variable | Mean (± SD) or N (%) | | MLE exponentiated $\beta$(95%CI) | | |
| --- | --- | --- | --- | --- | --- |
|  | office workers<br>(N = 84) | firefighters<br>(N= 86) | BDCPP | BCEP | DBuP |
| Age (years) | 48.3 (±10.5) | 47.5 (±4.6) | 1.00 (0.97,1.03) | 0.99 (0.94,1.04) | 1.00 (0.96,1.04) |
| Time lived in CA (years) | 35.5 (±14.5) | 40.0 (±10.1) | 0.99 (0.97,1.01) | 1.00 (0.97,1.04) | 1.01 (0.99,1.04) |
| U.S. Born | 62 (73.8%) | 77 (89.5%) | 0.80 (0.44,1.46) | 0.69 (0.24,2.02) | 0.90 (0.40,2.02) |
| **BMI <sup>b,c</sup>** |  |  |  |  |  |
| 18.5 – 24.9 | 43 (51.2%) | 33 (38.4%) | reference | reference | reference |
| 25.0 - 29.0 | 23 (27.4%) | 35 (40.7%) | 1.71 (1.02,2.84) | 2.70 (1.06,6.84) | 2.45 (1.28,4.68) |
| >30 | 16 (19.0%) | 13 (15.1%) | 0.84 (0.44,1.58) | 0.75 (0.23,2.50) | 1.55 (0.68,3.54) |
| **Race/ethnicity** |  |  |  |  |  |
| White | 37 (44.0%) | 40 (46.5%) | reference | reference | reference |
| Black | 5 (6.0%) | 9 (10.5%) | **2.52 (1.10,5.75)** | 0.76 (0.17,3.46) | 0.73 (0.25,2.17) |
| Latina | 13 (15.5%) | 19 (22.1%) | 0.56 (0.31,1.03) | 0.89 (0.30,2.67) | 0.82 (0.37,1.79) |
| Asian | 19 (22.6%) | 11 (12.8%) | 1.02 (0.55,1.88) | 1.60 (0.52,4.93) | 1.15 (0.50,2.63) |
| Other/multi | 10 (11.9%) | 7 (8.1%) | 0.66 (0.31,1.42) | 0.43 (0.10,1.89) | 0.29 (0.09,0.89) |
| **Education** |  |  |  |  |  |
| Some college or less | 15 (17.9%) | 48 (55.8%) | reference | reference | reference |
| Bachelors or greater | 69 (82.1%) | 38 (44.2%) | 1.65 (1.00,2.73) | 3.63 (1.49,8.84) | 2.24 (1.17,4.31) |

<sup>a</sup> Models adjusted for occupation (firefighter or office worker) and log(creatinine)
<sup>b</sup> CDC guidelines for BMI classification: normal weight 18.5-24.9; overweight 25.0-29.9, obese >30; BMI units: lb/in<sup>2</sup>
<sup>c</sup> seven participants declined to answer height weight questions

When we limited the analysis to firefighters, several variables were associated with increased FR levels, however none of these relationships were statistically significant (p-value ≤ 0.05) (**Table 3**). We observed that FR levels were higher firefighters who were on duty at the time of the sample collection (BDCPP exp(*β*) (95%CI) = 1.97 (0.91, 4.23) and BCEP (2.76 (0.91, 8.36)). We found slight associations by firefighters’ assigned role; officers and firefighters showed slightly elevated levels of BDCPP and BCEP compared to drivers. SCBA use in general was associated with lower FR levels with the exception of SCBA use during exterior fire suppression which was associated with higher mean BCEP compared to those who responded that they sometimes or less frequently did so. Firefighters assigned to one of the San Francisco Airport fire stations had elevated levels of BDCPP, BCEP, and DBuP. Fighting a fire within the 24 hours and 7 days prior to the sample collection was also slightly associated with BDCPP and BCEP levels. Use of firefighting foam, number of hours spent in vehicles per week for both home and work and the number of years worked with SFFD was not associated with FR levels.

**Table 3.** Adjusted^a^ proportional change in geometric mean of urine flame retardant concentration (ng/mL, DF >70%) by unit increase, or category change from the referent, of firefighter occupational activities and characteristics from individual MLE regression models

| Variable | Number responded | BDCPP | BCEP | DBuP |
| --- | --- | --- | --- | --- |
| Assigned to airport (yes) | 14 | 1.51 (0.66,3.44) | 1.73 (0.50,5.99) | 1.73 (0.69,4.33) |
| Years worked with SFFD | 86 | 0.93 (0.87,1.00) | 0.89 (0.80,1.00) | 0.92 (0.85,0.99) |
| Used firefighting foam past year (yes) | 25 | 0.99 (0.50,1.94) | 0.88 (0.32,2.45) | 0.80 (0.38,1.71) |
| On-duty at sample collection | 21 | 1.97 (0.91,4.23) | 2.76 (0.91,8.36) | 0.98 (0.41,2.33) |
| Hours spent in vehicle per week | 86 | 1.01 (0.98,1.04) | 1.04 (1.00,1.09) | 1.01 (0.98,1.05) |
| Fire in last 24hrs (yes) | 15 | 1.31 (0.58,2.94) | 1.52 (0.45,5.17) | 0.98 (0.40,2.43) |
| Fire in last 7 days (yes)<sup>b</sup> | 18 | 1.32 (0.61,2.89) | 1.86 (0.59,5.86) | 0.75 (0.30,1.90) |
| Fire in last month (yes)<sup>b</sup> | 44 | 1.24 (0.59,2.57) | 1.21 (0.40,3.66) | 0.95 (0.39,2.30) |
| **Position** |  |  |  |  |
| Driver | 21 | reference | reference | reference |
| Firefighter | 40 | 1.40 (0.65,3.01) | 1.32 (0.41,4.23) | 1.19 (0.50,2.83) |
| Officer | 25 | 1.62 (0.71,3.73) | 1.88 (0.53,6.70) | 1.40 (0.55,3.58) |
| **SCBA use with<sup>c</sup>** |  |  |  |  |
| Interior fire suppression (always vs often or less) | 60 | 0.75 (0.39,1.47) | 1.26 (0.45,3.48) | 0.95 (0.45,2.02) |
| Exterior fire suppression (often/always vs sometimes or less) | 32 | 0.90 (0.48,1.70) | 2.52 (0.92,6.88) | 1.01 (0.49,2.08) |
| Salvage & overhaul (often/always vs sometimes or less) | 26 | 0.51 (0.27,0.99) | 0.78 (0.28,2.16) | 0.50 (0.24,1.05) |

<sup>a</sup> Adjusted for log(creatinine)
<sup>b</sup> n = 66 firefighters who consented to giving researchers access to their SFFD fire history records;
<sup>c</sup> responses collected as 'never, rarely, sometimes, often, always' and were combined due to low frequency in individual response categories

### FR exposure and thyroid hormone levels

T_4_ and TSH were slightly negatively correlated with each other (Spearman correlation coefficient: −0.13, p-value = 0.1). We found that most participants were within the reference range (i.e. the range of levels that are considered normal)(Chiovato et al. 2019; Hollowell et al. 2002) for both TSH and T4 (**Table 4**). Only 6% (N = 11) of participants had TSH levels outside (below or above) of the reference range and sixteen participants (9%) had T4 levels outside the reference range, however the majority of those outside the reference range were firefighters (N = 14).

**Table 4.** Geometric mean, geometric standard deviation and select percentiles for thyroid hormones TSH and T_4_ in WFBC study participants N = 165 Percentiles

| Hormone | Reference range<sup>a</sup> | GM (GSD) | Min | Max | Percentiles |  |  |  |
| --- | --- | --- | --- | --- | --- | --- | --- | --- |
|  |  |  |  |  | 25% | 50% | 75% | 95% |
| TSH | 0.4 to 4.0 mIU/L | 1.26 (2.06) | 0.03 | 11.4 | 0.88 | 1.36 | 1.96 | 3.36 |
| T4 | 4.6 to 12 ug/dL | 6.41 (1.30) | 3.69 | 20.58 | 5.43 | 6.36 | 7.26 | 9.29 |

Abbreviations: GM = Geometric mean, GSD = geometric Standard deviation.
<sup>a</sup>Chiovato et al. 2019; Hollowell et al. 2002

TSH levels were not significantly different between firefighters and office workers (permutation test p-value = 0.6) however T_4_ levels did vary slightly by occupation (permutation test p-value = 0.06). We assessed the relationship of potential confounders with TSH and T_4_ as a consideration for inclusion into OLS regression models. Since neither race/ethnicity nor education were associated with TSH or T_4_ in our data (data not shown), we did not include them in final models. Although age was not associated with TH in our study population, it was included as a potential confounder based on prior literature indicating it may be an important factor for determining TH levels (Hollowell et al. 2002).

We applied OLS regression models to assess the impact of FR levels on T_4_ and TSH levels controlling for log(creatinine) and age. FR levels were not associated with TSH in our models (**Table S2**); however, we did see a relationship between several FR and T_4_ levels. BDCPP, which we defined as continuous (DF>70%) in OLS regression models was negatively associated with T_4_ levels (**Table 5**). In the full cohort, a two-fold increase in BDCPP levels was associated with a decrease of 1.95% (95% CI 3.57, 0.29) in T_4_, and in models limited to firefighters, a two-fold increase in BDCCP was associated with a T_4_ decrease of 2.88% (5.278, 0.417), controlling for age and log(creatine). The percent change among office workers was smaller and not significant.

**Table 5.** β(95% confidence interval (CI)) and percent change (95% CI) in T4 levels for each doubling of BDCPP in adjusted ^a^ OLS regression models

| **Model** | **$\beta$(95%CI)** | **Percent change (95%CI)** |
| --- | --- | --- |
| Full cohort | -0.028 (-0.053,-0.004) | -1.95 (-3.57,-0.29) |
| Firefighters only | -0.042 (-0.078,-0.006) | -2.88 (-5.28,-0.42) |
| Office workers only | 0.003 (-0.036,0.043) | 0.23 (-2.49,3.026) |

<sup>a</sup>Models adjusted for age and log(creatinine); Values below LOD replaced with LOD/sqrt(2)

Due to large differences in DF for DBuP, BCEP, TBBPA and DPCP between firefighters and office workers, we ran separate OLS regression models by occupation for each FR as a predictor and T_4_ as a continuous outcome. For office workers, we categorized DBuP, BCEP, TBBPA and DPCP into <LOD, ≥ LOD. Among firefighters we categorized TBBPA and DPCP as <LOD, ≥LOD and categorized DBuP and BCEP as <LOD, LOD to 50^th^ %, >50^th^ %. Because the DF for DBuP and BCEP was >70% for firefighters we also ran OLS regression models with the continuous DBuP and BCEP. Among firefighters, DBuP was slightly associated with a decrease in T_4_ levels among those at the LOD or above compared to below the LOD. DBuP was not significantly associated with T_4_ among office workers nor were BCEP, TBBPA and DPCP associated with T_4_ levels in either firefighters or office workers (**Table 6**).

**Table 6.** Percent change (95% CI) in T4 for each category increase in FR compared to <LOD for firefighters and office workers from adjusted OLS regression models^a^

| Firefighters |  | Office Workers |  |
| --- | --- | --- | --- |
| DBuP |  |  |  |
| <LOD | reference | <LOD | reference |
| LOD-50th% | -6.07 (-19.45,9.53) | >LOD | 5.57 (-6.21,18.83) |
| >50th% | -12.68 (-24.94,1.59) |  |  |
| *Continuous* | -2.01 (-4.63,0.68) |  |  |
| BCEP |  |  |  |
| <LOD | reference | <LOD | reference |
| LOD-50th % | 8.05 (-7.13,21.61) | >LOD | -1.16 (-11.59,10.49) |
| >50th% | 5.23 (-1.79,0.61) |  |  |
| *Continuous* | 0.623 (-1.58,2.88) |  |  |
| TBBPA |  |  |  |
| <LOD | reference | <LOD | reference |
| >LOD | 4.30 (-6.29,16.08) | >LOD | 6.28 (-4.71,18.54) |
| DPCP<sup>b</sup> |  |  |  |
| <LOD | reference |  |  |
| >LOD | -9.21 (-19.29,2.14) |  |  |

<sup>a</sup>Models adjusted age and log(creatinine); If DF < 50% OPFR values were categorized into <= LOD or >LOD; those OPFRs with a DF of at least 50% were categorized as <=LOD, >LOD to 50th, and >50th; DBuP and BCEP DF >70 %
<sup>b</sup> detection frequency among office workers <25%

## Discussion

To our knowledge this is the first study to measure exposures to replacement FR and effects in thyroid function among women firefighters compared to office workers. In addition, this paper confirms prior reports that firefighters have higher levels of OPFRs compared to the general population and contributes further evidence that OPFRs are an endocrine disrupting compound.

### Flame retardant levels and predictors of exposure

FR were detected in all firefighters tested and most office workers. Women firefighters had higher detection frequencies and higher average levels of flame retardants compared to office workers. BDCPP, BCEP, and DBuP were detected more frequently and at higher concentrations among firefighters compared to office workers. BDCPP is a metabolite of TDCPP, a FR that is commonly detected in household dust and in furniture foam (Hammel et al. 2017), which may help explain why office workers also had a high detection frequency of this compound. In fact, studies of adult women (age 18+) and children (ages 6-12) from the 2013-2014 National Health and Nutrition Examination Survey (NHANES), a nationally representative sample of the US population, had similar levels to the office workers of our study (Ospina et al. 2018).

Few demographic variables were associated with FR levels. Age, for example was not associated with FR in our study; Conversely, the positive relationship between OFPR exposures and BMI is consistent with other studies (Boyle et al. 2019). While Black participants had higher levels of BDCPP compared to White participants when controlling for age and occupation, this relationship may in fact reflect other unknown exposures and be a function of the limited numbers of Black participants in both the firefighters and office worker groups.

California’s flammability standards and the changes implemented in 2013 may have impacted the levels of chemicals that we observed in our study population. Concerns about the bioaccumulative properties of brominated flame retardants and the updated regulation TB177-2013, led to a shift in the types of FR used in furniture. Indeed, studies have observed a decrease in PBDEs in people and the environment, while levels of OPFRs and other replacement flame retardants are increasing in dust and human biomonitoring studies (Dodson et al. 2012; Ospina et al. 2018; Stapleton et al. 2008; Zota et al. 2013). The half-lives of these replacement chemicals are not known, though they are generally considered to be shorter than that of PBDEs and potentially on the order of hours or days (Carignan et al. 2013; Nomeir et al. 1981). However, furniture in homes and offices can be a reservoir for both legacy and replacement FR; furniture is not frequently replaced and the chemicals applied to them are relatively stable enabling FR to persist in homes and offices for decades (Zota et al. 2013). The elevated levels of BDCPP and other flame retardants in furniture, homes, and offices, may also be an important exposure source among firefighters as well as the office workers in our study. Firefighter turnout gear and protective equipment is plausibly another source of FR for firefighters, since they have been shown to be treated with flame retardant chemicals. Similarly, FR-containing furniture may add to the toxic burden of fighting fires and translate to higher exposures among firefighters when they respond to fires. Studies have shown that fighting fires can contaminate firefighter gear, trucks, engines and equipment, bringing chemical exposures indoors (Banks et al. 2020; Mayer et al. 2019; Shen et al. 2018).

Fire station dust is potentially an important source of FR exposure among firefighters. The increased levels of OPFR found in the women firefighters compared to office workers is consistent with studies measuring elevated levels of OPFRs in fire station dust in the U.S. and abroad. Shen *et al*. (2018) found higher levels of TDCPP, tri-n-butyl phosphate (TNBP), and tris(2-chloroisopropyl)phosphate (TCPP), in dust collected in 2015 from fire stations across the U.S., compared to levels measured in homes and other occupational settings (Shen et al. 2018). Similarly, a study of Australian fire station dust found higher median OPFR levels in fire stations than from dust samples collected in homes and offices (Banks et al. 2020). Firefighters who were on duty at the time of the sample collection had higher levels of two FR chemicals supporting evidence that being at work and potentially in the fire station increases their exposures. We expect that firefighter gear may also be a source of exposure; however, we do not have information on the age, type, manufacturer or chemical composition of turnout gear that firefighters use in San Francisco. A 2016 study of firefighter gear found that some new hoods and gloves had detectable levels of brominated flame retardants (Alexander and Baxter 2016) but to our knowledge studies have not analyzed gear for OPFRs. Nevertheless, firefighter gear may still contain FR compounds that can additionally contribute to exposures from dust accumulated on the gear from fires and calls (Alexander and Baxter 2016). A recent study of firefighting gear found that fighting fires may contaminate the hoods firefighters wear under their protective equipment with OPFRs (Mayer et al. 2019). Additionally, laundering may have a limited impact on reducing exposures and, in some cases, may cross-contaminate gear (Mayer et al. 2019).

### FR exposure and thyroid hormone levels

The toxicity of most replacement chemical flame retardants has not been fully characterized. One of the few FRs that has been assessed for toxicity and adverse health effects is TDCPP which, due to its potential for carcinogenicity has been included in California’s Prop 65 list (Office of Environmental Health Hazard Assessment 2015). In fact TDCPP, which has been used as a FR replacement in PUF and fabrics, is structurally similar to tris(2,3-dibromopropyl) phosphate (TDBPP or brominated “Tris”) which was banned in the 1970’s from use in children’s pajamas because of its mutagenic and carcinogenic properties (Dodson et al. 2012).

Our study demonstrated that firefighters in San Francisco have higher levels of FR than office workers and contributes to the limited literature of thyroid hormone disruption in women in relation to FR exposure. This association of FR with decreased T_4_ was primarily observed among firefighters, and particularly for BDCPP, which had the highest DF of the FR we measured. The broader range and much higher levels of exposure among the firefighters in our cohort may have allowed us to see an effect of FR exposure on T_4_ that we did not observe among office workers.

*In vitro* studies provide evidence for the mechanistic plausibility of TH disruption from OPFR exposures. However, the mechanism of effect is not yet fully understood. Hill et al. (2018) tested the competition of binding of T_4_ and TTR (one of many transport proteins for T_4_). They found that TDCPP and other organophosphate tri- and di-esters increased the binding affinity of T_4_ with TTR. They hypothesized that organophosphate compounds may bind to the surface of TTR, creating a conformational change allowing more T_4_ binding and increasing the delivery of T_4_ to target cells resulting in lower circulating levels of T_4_ and disrupting T_4_ homeostasis (Hill et al. 2018). Our study provides evidence that exposure to TDCPP and, to a lesser extent, DBuP exposure can affect TH levels. While our study does not have a clinical outcome, and the changes we observed were relatively small, even small disruptions to TH can have multiple adverse downstream health effects (Boas et al. 2012) even within normal ranges (Taylor et al. 2013). Thyroid dysfunction affects up to 5% of the population and is more likely to affect women than men (Hollowell et al. 2002). Over the past two decades the incidence of thyroid cancer has also increased and cannot be fully explained by improved testing and diagnosis (Ward et al. 2010). Identifying environmental chemical exposures that may be associated with biological changes, such as thyroid disruption could be relevant to adverse health outcomes such as cardiovascular disease (Rodondi et al. 2006) and thyroid disease (Ward et al. 2010), brain development of offspring during gestation, and long-term adverse health outcomes such as cancer (Krashin et al. 2019).

We were unable to identify specific sources of FRs or why firefighters had higher levels of exposure than office workers. While durable consumer goods such as couches and mattresses may contribute to levels in the general population and fire station dust may be an important source in firefighters, this does not fully explain where the FR exposures are coming from and why the levels are so much higher in fire fighters. Future studies need to elucidate potential sources of exposure to legacy and replacement FR which would facilitate the development and promotion of effective exposure prevention strategies.

### Limitations

This was a cross sectional study of FR exposure and effects on thyroid function, which precludes making inferences regarding causality. Nevertheless, our findings confirm an association between TH levels and TDCPP, one that has also been identified in studies conducted *in vitro, in vivo* and in limited human studies (Farhat et al. 2013; Meeker and Stapleton 2010; Wang et al. 2013a).

While we did not detect PBDE metabolites in our study population, analyzing these metabolites in urine is insensitive, and therefore these results were somewhat expected. OH-BDE metabolites are more commonly measured in serum and these chemicals have been found in participants of other studies conducted during the same time as WFBC study (Park et al. 2015; Parry et al. 2018). Therefore, although we did not find OH-BDE metabolites in any of our samples, this may be due to the type of biological matrix we measured rather than an absence of exposure. Another potential limitation is that we used a mix of morning void and spot urine samples when measuring FR levels, potentially increasing the variability of the urine concentration and of chemicals measured in our samples. However, morning void or spot samples were selected at random for analysis, therefore the variability would be non-differential between firefighter and office workers and not likely to affect the differences we observed in their chemical levels nor the FR’s relationship with potential covariates. In addition, we measured and adjusted for creatinine to account for urine dilution and reduce some of this variability (Barr et al. 2005).

Thyroid disease and taking thyroid hormone replacement medications may alter the TH levels. We asked participants “do you take any hormones other than birth control, such as premarin? if so what?” Some participants disclosed taking TH medications or having thyroid problems, however, the question may not have accurately captured all the participants with thyroid dysfunction or those who take TH replacements. Therefore, we may not have excluded everyone with artificial or abnormal TH levels due to illness or medication rather than chemical exposure. This could over- or under-estimate thyroid hormone levels depending on the thyroid problem, and we would not be able to predict in what direction this could affect our results. Finally, we may also have had limited statistical power, due to our modest sample size, to assess the relationship between FR levels and predictors of exposure. This may have precluded our ability to see the full effect of FR exposure on TH disruption, especially at the lower exposure levels we observed in office workers.

## Conclusion

This is the first study to measure flame retardant exposure in a cohort of women firefighters. Most participants had detectable levels of at least one organophosphate flame retardant, and both detection frequencies and levels were much higher among women firefighters than in women office workers; Median levels of BDCPP were five times higher in firefighters than for office workers. Additionally, exposure to BDCPP, and to a smaller extent DBuP, was associated with decreased levels of thyroxine particularly among firefighters—a twofold increase in BDCPP was associated with a 2.88% decrease in T_4._ The observed thyroid hormone disruption may indicate potential biological perturbations resulting from occupational exposure to these flame retardants. Further research is needed to understand sources in occupational settings and fully characterize potential health impacts of these replacement flame retardants.

## Data Availability

Our IRB and human subjects protection precludes our ability to share data.

## Acknowledgements

The authors thank all of the WFBC study participants for their contribution to the study. This work is supported by the California Breast Cancer Research Program #19BB-2900 and #23BB-1700 & 1701 & 1702 (JT, RG, CC, MM, CA, RAR, HB, VB, RMF), the National Institute of Environmental Health Sciences R01ES027051 (RMF), the National Institute for Occupational Safety and Health, Targeted Research Training Program T42 OH008429 (JT), the San Francisco Firefighter Cancer Prevention Foundation (HB) and the International Association of Firefighters-Local 798. We thank Anthony Stefani, Emily O’Rourke, Nancy Carmona, Karen Kerr, Julie Mau, Natasha Parks, Lisa Holdcroft, San Francisco Fire Chief Jeanine Nicholson, former San Francisco Fire Chief Joanne Hayes-White, Sharyle Patton, Connie Engel and Nancy Buermeyer for their contributions to the study. RAR and VB, are employed at the Silent Spring Institute, a scientific research organization dedicated to studying environmental factors in women’s health. The Institute is a 501(c)3 public charity funded by federal grants and contracts, foundation grants, and private donations, including from breast cancer organizations. HB is former president and member of United Fire Service Women, a 501(c)3 public charity dedicated to supporting the welfare of women in the San Francisco Fire Department. The authors declare they have no actual or potential competing financial interests.

## References

Alexander BM, Baxter CS. 2016. Flame-retardant contamination of firefighter personal protective clothing - A potential health risk for firefighters. J Occup Environ Hyg 13:D148–155; doi:10.1080/15459624.2016.1183016.

Antibodies-online. 2020. T3, T4, TSH ELISA Kit - Product details.

Banks APW, Engelsman M, He C, Wang X, Mueller JF. 2020. The occurrence of PAHs and flame-retardants in air and dust from Australian fire stations. J Occup Environ Hyg 17:73–84; doi:10.1080/15459624.2019.1699246.

Barr DB, Wilder LC, Caudill SP, Gonzalez AJ, Needham LL, Pirkle JL. 2005. Urinary Creatinine Concentrations in the U.S. Population: Implications for Urinary Biologic Monitoring Measurements. Environ Health Perspect 113:192–200; doi:10.1289/ehp.7337.

Birnbaum LS, Staskal DF. 2004. Brominated flame retardants: cause for concern? Environ Health Perspect 112:9–17; doi:10.1289/ehp.6559.

Boas M, Feldt-Rasmussen U, Main KM. 2012. Thyroid effects of endocrine disrupting chemicals. Mol Cell Endocrinol 355:240–248; doi:10.1016/j.mce.2011.09.005.

Boyle M, Buckley JP, Quiros-Alcala L. 2019. Associations between urinary organophosphate ester metabolites and measures of adiposity among U.S. children and adults: NHANES 2013-2014. Environ Int 127:754–763; doi:10.1016/j.envint.2019.03.055.

Brown FR, Whitehead TP, Park J-S, Metayer C, Petreas MX. 2014. Levels of non-polybrominated diphenyl ether brominated flame retardants in residential house dust samples and fire station dust samples in California. Environmental Research 135:9–14; doi:10.1016/j.envres.2014.08.022.

Bureau of Electronic and Appliance. 2018. Technical Bulletin 117-2013. Available: http://www.bearhfti.ca.gov/about_us/tb117_2013.pdf [accessed 3 March 2018].

Calsolaro V, Pasqualetti G, Niccolai F, Caraccio N, Monzani F. 2017. Thyroid Disrupting Chemicals. Int J Mol Sci 18; doi:10.3390/ijms18122583.

Carignan CC, McClean MD, Cooper EM, Watkins DJ, Fraser AJ, Heiger-Bernays W, Stapleton HM, Webster TF. 2013. Predictors of Tris(1,3-dichloro-2-propyl) phosphate Metabolite in the Urine of Office Workers. Environ Int 55:56–61; doi:10.1016/j.envint.2013.02.004.

Castorina R, Bradman A, Stapleton HM, Butt C, Avery D, Harley KG, Gunier RB, Holland N, Eskenazi B. 2017. Current-use flame retardants: Maternal exposure and neurodevelopment in children of the CHAMACOS cohort. Chemosphere 189:574–580; doi:10.1016/j.chemosphere.2017.09.037.

Caux C, O’Brien C, Viau C. 2002. Determination of firefighter exposure to polycyclic aromatic hydrocarbons and benzene during fire fighting using measurement of biological indicators. Appl Occup Environ Hyg 17:379–386; doi:10.1080/10473220252864987.

Chiovato L, Magri F, Carlé A. 2019. Hypothyroidism in Context: Where We’ve Been and Where We’re Going. Adv Ther 36:47–58; doi:10.1007/s12325-019-01080-8.

Covaci A, Harrad S, Abdallah MA-E, Ali N, Law RJ, Herzke D, de Wit CA. 2011. Novel brominated flame retardants: A review of their analysis, environmental fate and behaviour. Environment International 37:532–556; doi:10.1016/j.envint.2010.11.007.

Daniels RD, Bertke S, Dahm MM, Yiin JH, Kubale TL, Hales TR, Baris D, Zahm SH, Beaumont JJ, Waters KM, Pinkerton LE. 2015. Exposure-response relationships for select cancer and non-cancer health outcomes in a cohort of U.S. firefighters from San Francisco, Chicago and Philadelphia (1950-2009). Occup Environ Med 72:699–706; doi:10.1136/oemed-2014-102671.

Daniels RD, Kubale TL, Yiin JH, Dahm MM, Hales TR, Baris D, Zahm SH, Beaumont JJ, Waters KM, Pinkerton LE. 2014. Mortality and cancer incidence in a pooled cohort of US firefighters from San Francisco, Chicago and Philadelphia (1950-2009). Occup Environ Med 71:388–397; doi:10.1136/oemed-2013-101662.

Diamanti-Kandarakis E, Bourguignon J-P, Giudice LC, Hauser R, Prins GS, Soto AM, Zoeller RT, Gore AC. 2009. Endocrine-disrupting chemicals: an Endocrine Society scientific statement. Endocr Rev 30:293–342; doi:10.1210/er.2009-0002.

Dishaw LV, Macaulay LJ, Roberts SC, Stapleton HM. 2014. Exposures, mechanisms, and impacts of endocrine-active flame retardants. Curr Opin Pharmacol 19:125–133; doi:10.1016/j.coph.2014.09.018.

Dobraca D, Israel L, McNeel S, Voss R, Wang M, Gajek R, Park J-S, Harwani S, Barley F, She J, Das R. 2015. Biomonitoring in California firefighters: metals and perfluorinated chemicals. J Occup Environ Med 57:88–97; doi:10.1097/JOM.0000000000000307.

Dodson RE, Perovich LJ, Covaci A, Van den Eede N, Ionas AC, Dirtu AC, Brody JG, Rudel RA. 2012. After the PBDE phase-out: a broad suite of flame retardants in repeat house dust samples from California. Environ Sci Technol 46:13056–13066; doi:10.1021/es303879n.

Dodson RE, Van den Eede N, Covaci A, Perovich LJ, Brody JG, Rudel RA. 2014. Urinary biomonitoring of phosphate flame retardants: levels in California adults and recommendations for future studies. Environ Sci Technol 48:13625–13633; doi:10.1021/es503445c.

Farhat A, Crump D, Chiu S, Williams KL, Letcher RJ, Gauthier LT, Kennedy SW. 2013. In Ovo effects of two organophosphate flame retardants--TCPP and TDCPP--on pipping success, development, mRNA expression, and thyroid hormone levels in chicken embryos. Toxicol Sci 134:92–102; doi:10.1093/toxsci/kft100.

Fent KW, Eisenberg J, Snawder J, Sammons D, Pleil JD, Stiegel MA, Mueller C, Horn GP, Dalton J. 2014. Systemic exposure to PAHs and benzene in firefighters suppressing controlled structure fires. Ann Occup Hyg 58:830–845; doi:10.1093/annhyg/meu036.

Gore AC, Chappell VA, Fenton SE, Flaws JA, Nadal A, Prins GS, Toppari J, Zoeller RT. 2015. EDC-2: The Endocrine Society’s Second Scientific Statement on Endocrine-Disrupting Chemicals. Endocr Rev 36:E1–E150; doi:10.1210/er.2015-1010.

Grashow R, Bessonneau V, Gerona RR, Wang A, Trowbridge J, Lin T, Buren H, Rudel RA, Morello-Frosch R. 2020. Integrating exposure knowledge and serum suspect screening as a new approach to biomonitoring: An application in firefighters and office workers. Environ Sci Technol; doi:10.1021/acs.est.9b04579.

Gravel S, Lavoue J, Bakhiyi B, Lavoie J, Roberge B, Patry L, Bouchard MF, Verner M-A, Zayed J, Labreche F. 2020. Multi-exposures to suspected endocrine disruptors in electronic waste recycling workers: Associations with thyroid and reproductive hormones. Int J Hyg Environ Health 225:113445; doi:10.1016/j.ijheh.2019.113445.

Hammel SC, Hoffman K, Lorenzo AM, Chen A, Phillips AL, Butt CM, Sosa JA, Webster TF, Stapleton HM. 2017. Associations Between Flame Retardant Applications in Furniture Foam, House Dust Levels, and Residents’ Serum Levels. Environ Int 107:181–189; doi:10.1016/j.envint.2017.07.015.

Hill KL, Hamers T, Kamstra JH, Willmore WG, Letcher RJ. 2018. Organophosphate triesters and selected metabolites enhance binding of thyroxine to human transthyretin in vitro. Toxicol Lett 285:87–93; doi:10.1016/j.toxlet.2017.12.030.

Hoffman K, Fang M, Horman B, Patisaul HB, Garantziotis S, Birnbaum LS, Stapleton HM. 2014. Urinary tetrabromobenzoic acid (TBBA) as a biomarker of exposure to the flame retardant mixture Firemaster® 550. Environ Health Perspect 122:963–969; doi:10.1289/ehp.1308028.

Hollowell JG, Staehling NW, Flanders WD, Hannon WH, Gunter EW, Spencer CA, Braverman LE. 2002. Serum TSH, T(4), and thyroid antibodies in the United States population (1988 to 1994): National Health and Nutrition Examination Survey (NHANES III). J Clin Endocrinol Metab 87:489–499; doi:10.1210/jcem.87.2.8182.

Jin C, Sun Y, Islam A, Qian Y, Ducatman A. 2011. Perfluoroalkyl acids including perfluorooctane sulfonate and perfluorohexane sulfonate in firefighters. J Occup Environ Med 53:324–328; doi:10.1097/JOM.0b013e31820d1314.

Kim H, Rebholz CM, Wong E, Buckley JP. 2020. Urinary organophosphate ester concentrations in relation to ultra-processed food consumption in the general US population. Environmental Research 182:109070; doi:10.1016/j.envres.2019.109070.

Knudsen N, Laurberg P, Rasmussen LB, Bülow I, Perrild H, Ovesen L, Jørgensen T. 2005. Small Differences in Thyroid Function May Be Important for Body Mass Index and the Occurrence of Obesity in the Population. J Clin Endocrinol Metab 90:4019–4024; doi:10.1210/jc.2004-2225.

Krashin E, Piekiełko-Witkowska A, Ellis M, Ashur-Fabian O. 2019. Thyroid Hormones and Cancer: A Comprehensive Review of Preclinical and Clinical Studies. Front Endocrinol (Lausanne) 10; doi:10.3389/fendo.2019.00059.

Laitinen JA, Koponen J, Koikkalainen J, Kiviranta H. 2014. Firefighters’ exposure to perfluoroalkyl acids and 2-butoxyethanol present in firefighting foams. Toxicol Lett 231:227–232; doi:10.1016/j.toxlet.2014.09.007.

Lee DJ, Koru-Sengul T, Hernandez MN, Caban-Martinez AJ, McClure LA, Mackinnon JA, Kobetz EN. 2020. Cancer risk among career male and female Florida firefighters: Evidence from the Florida Firefighter Cancer Registry (1981-2014). Am J Ind Med 63:285–299; doi:10.1002/ajim.23086.

LeMasters GK, Genaidy AM, Succop P, Deddens J, Sobeih T, Barriera-Viruet H, Dunning K, Lockey J. 2006. Cancer risk among firefighters: a review and meta-analysis of 32 studies. J Occup Environ Med 48:1189–1202; doi:10.1097/01.jom.0000246229.68697.90.

Lerro CC, Koutros S, Andreotti G, Friesen MC, Alavanja MC, Blair A, Hoppin JA, Sandler DP, Lubin JH, Ma X, Zhang Y, Beane Freeman LE. 2015. Organophosphate insecticide use and cancer incidence among spouses of pesticide applicators in the Agricultural Health Study. Occup Environ Med 72:736–744; doi:10.1136/oemed-2014-102798.

Ma F, Fleming LE, Lee DJ, Trapido E, Gerace TA. 2006. Cancer incidence in Florida professional firefighters, 1981 to 1999. J Occup Environ Med 48:883–888; doi:10.1097/01.jom.0000235862.12518.04.

Mayer AC, Fent KW, Bertke S, Horn GP, Smith DL, Kerber S, La Guardia MJ. 2019. Firefighter hood contamination: Efficiency of laundering to remove PAHs and FRs. J Occup Environ Hyg 16:129–140; doi:10.1080/15459624.2018.1540877.

Meeker JD, Stapleton HM. 2010. House dust concentrations of organophosphate flame retardants in relation to hormone levels and semen quality parameters. Environ Health Perspect 118:318–323; doi:10.1289/ehp.0901332.

Nomeir AA, Kato S, Matthews HB. 1981. The metabolism and disposition of tris(1,3-dichloro-2-propyl) phosphate (Fyrol FR-2) in the rat. Toxicol Appl Pharmacol 57:401–413; doi:10.1016/0041-008x(81)90238-6.

Office of Environmental Health Hazard Assessment. 2015. Tris(1,3-dichloro-2-propyl) Phosphate (TDCPP). OEHHA. Available: https://oehha.ca.gov/proposition-65/chemicals/tris13-dichloro-2-propyl-phosphate-tdcpp [accessed 25 April 2020].

Ospina M, Jayatilaka NK, Wong L-Y, Restrepo P, Calafat AM. 2018. Exposure to organophosphate flame retardant chemicals in the U.S. general population: Data from the 2013-2014 National Health and Nutrition Examination Survey. Environ Int 110:32–41; doi:10.1016/j.envint.2017.10.001.

Park J-S, Voss RW, McNeel S, Wu N, Guo T, Wang Y, Israel L, Das R, Petreas M. 2015. High Exposure of California Firefighters to Polybrominated Diphenyl Ethers. Environmental Science & Technology 49:2948–2958; doi:10.1021/es5055918.

Parry E, Zota AR, Park J-S, Woodruff TJ. 2018. Polybrominated diphenyl ethers (PBDEs) and hydroxylated PBDE metabolites (OH-PBDEs): A six-year temporal trend in Northern California pregnant women. Chemosphere 195:777–783; doi:10.1016/j.chemosphere.2017.12.065.

Pinkerton L, Bertke SJ, Yiin J, Dahm M, Kubale T, Hales T, Purdue M, Beaumont JJ, Daniels R. 2020. Mortality in a cohort of US firefighters from San Francisco, Chicago and Philadelphia: an update. Occup Environ Med 77:84–93; doi:10.1136/oemed-2019-105962.

Preston EV, McClean MD, Claus Henn B, Stapleton HM, Braverman LE, Pearce EN, Makey CM, Webster TF. 2017. Associations between urinary diphenyl phosphate and thyroid function. Environ Int 101:158–164; doi:10.1016/j.envint.2017.01.020.

R Core Team. 2015. R: A language and environment for statistical computing. R Foundation for Statistical Computing:Vienna, Austria.

Rodgers KM, Udesky JO, Rudel RA, Brody JG. 2018. Environmental chemicals and breast cancer: An updated review of epidemiological literature informed by biological mechanisms. Environ Res 160:152–182; doi:10.1016/j.envres.2017.08.045.

Rodondi N, Aujesky D, Vittinghoff E, Cornuz J, Bauer DC. 2006. Subclinical hypothyroidism and the risk of coronary heart disease: a meta-analysis. Am J Med 119:541–551; doi:10.1016/j.amjmed.2005.09.028.

RStudio Team. 2016. RStudio: Integrated Development for R. RStudio Inc.:Boston, MA.

Rudel RA, Ackerman JM, Attfield KR, Brody JG. 2014. New exposure biomarkers as tools for breast cancer epidemiology, biomonitoring, and prevention: a systematic approach based on animal evidence. Environ Health Perspect 122:881–895; doi:10.1289/ehp.1307455.

Rudel RA, Fenton SE, Ackerman JM, Euling SY, Makris SL. 2011. Environmental exposures and mammary gland development: state of the science, public health implications, and research recommendations. Environ Health Perspect 119:1053–1061; doi:10.1289/ehp.1002864.

Schindler BK, Weiss T, Schütze A, Koslitz S, Broding HC, Bünger J, Brüning T. 2013. Occupational exposure of air crews to tricresyl phosphate isomers and organophosphate flame retardants after fume events. Arch Toxicol 87:645–648; doi:10.1007/s00204-012-0978-0.

Shaw SD, Berger ML, Harris JH, Yun SH, Wu Q, Liao C, Blum A, Stefani A, Kannan K. 2013. Persistent organic pollutants including polychlorinated and polybrominated dibenzo-p-dioxins and dibenzofurans in firefighters from Northern California. Chemosphere 91:1386–1394; doi:10.1016/j.chemosphere.2012.12.070.

Shen B, Whitehead TP, Gill R, Dhaliwal J, Brown FR, Petreas M, Patton S, Hammond SK. 2018. Organophosphate flame retardants in dust collected from United States fire stations. Environment International 112:41–48; doi:10.1016/j.envint.2017.12.009.

Shen B, Whitehead TP, McNeel S, Brown FR, Dhaliwal J, Das R, Israel L, Park J-S, Petreas M. 2015. High levels of polybrominated diphenyl ethers in vacuum cleaner dust from California fire stations. Environ Sci Technol 49:4988–4994; doi:10.1021/es505463g.

Stapleton HM, Allen JG, Kelly SM, Konstantinov A, Klosterhaus S, Watkins D, McClean MD, Webster TF. 2008. Alternate and new brominated flame retardants detected in U.S. house dust. Environ Sci Technol 42:6910–6916; doi:10.1021/es801070p.

Stapleton HM, Sharma S, Getzinger G, Ferguson PL, Gabriel M, Webster TF, Blum A. 2012. Novel and high volume use flame retardants in US couches reflective of the 2005 PentaBDE phase out. Environ Sci Technol 46:13432–13439; doi:10.1021/es303471d.

Taylor PN, Razvi S, Pearce SH, Dayan CM. 2013. Clinical review: A review of the clinical consequences of variation in thyroid function within the reference range. J Clin Endocrinol Metab 98:3562–3571; doi:10.1210/jc.2013-1315.

Trowbridge J, Gerona RR, Lin T, Rudel RA, Bessonneau V, Buren H, Morello-Frosch R. 2020. Exposure to Perfluoroalkyl Substances in a Cohort of Women Firefighters and Office Workers in San Francisco. Environ Sci Technol 54:3363–3374; doi:10.1021/acs.est.9b05490.

Van den Eede N, Dirtu AC, Ali N, Neels H, Covaci A. 2012. Multi-residue method for the determination of brominated and organophosphate flame retardants in indoor dust. Talanta 89:292–300; doi:10.1016/j.talanta.2011.12.031.

van der Veen I, de Boer J. 2012a. Phosphorus flame retardants: properties, production, environmental occurrence, toxicity and analysis. Chemosphere 88:1119–1153; doi:10.1016/j.chemosphere.2012.03.067.

van der Veen I, de Boer J. 2012b. Phosphorus flame retardants: Properties, production, environmental occurrence, toxicity and analysis. Chemosphere 88:1119–1153; doi:10.1016/j.chemosphere.2012.03.067.

Wang Q, Liang K, Liu J, Yang L, Guo Y, Liu C, Zhou B. 2013a. Exposure of zebrafish embryos/larvae to TDCPP alters concentrations of thyroid hormones and transcriptions of genes involved in the hypothalamic-pituitary-thyroid axis. Aquat Toxicol 126:207–213; doi:10.1016/j.aquatox.2012.11.009.

Wang Y, Starling AP, Haug LS, Eggesbo M, Becher G, Thomsen C, Travlos G, King D, Hoppin JA, Rogan WJ, Longnecker MP. 2013b. Association between Perfluoroalkyl substances and thyroid stimulating hormone among pregnant women: a cross-sectional study. Environ Health 12:76; doi:10.1186/1476-069X-12-76.

Ward EM, Jemal A, Chen A. 2010. Increasing incidence of thyroid cancer: is diagnostic scrutiny the sole explanation? Future Oncology 6:185–188; doi:10.2217/fon.09.161.

Zoeller RT, Tan SW, Tyl RW. 2007. General background on the hypothalamic-pituitary-thyroid (HPT) axis. Crit Rev Toxicol 37:11–53; doi:10.1080/10408440601123446.

Zota AR, Linderholm L, Park J-S, Petreas M, Guo T, Privalsky ML, Zoeller RT, Woodruff TJ. 2013. Temporal Comparison of PBDEs, OH-PBDEs, PCBs, and OH-PCBs in the Serum of Second Trimester Pregnant Women Recruited from San Francisco General Hospital, California. Environ Sci Technol 47:11776–11784; doi:10.1021/es402204y.

