## Supplemental tables and figures for "Organophosphate and organohalogen flame-retardant exposure and thyroid hormone disruption in a cohort of female firefighters and office workers from San Francisco"

**Supplemental Information**

| **Table S1**. Descriptive statistics and percentiles of flame-retardant^a^ measurements for firefighters (n= 86) and office workers (n= 84) | | | | | | | | |
| --- | --- | --- | --- | --- | --- | --- | --- | --- |
|  |  |  |  |  | Percentiles | | | |
| **Chemical** | **Group** | **LOD** | **DF%** | **GM (GSD)** | **25th** | **50th** | **75th** | **95th** |
| **BCEP** |  |  |  |  |  |  |  |  |
|  | Firefighter | 0.1 | 78 | 0.85 (5.74) | 0.23 | 1.22 | 3.98 | 8.51 |
|  | Office worker | 0.1 | 39 | <LOD | <LOD | <LOD | 0.485 | 3.31 |
| **BDCPP** |  |  |  |  |  |  |  |  |
|  | Firefighter | 0.2 | 100 | 4.08 (4.53) | 1.30 | 5.37 | 13.3 | 32.22 |
|  | Office worker | 0.2 | 90 | 0.96 (3.99) | <LOD | 0.92 | 2.3425 | 8.72 |
| **DBuP** |  |  |  |  |  |  |  |  |
|  | Firefighter | 0.1 | 83 | 0.41 (3.92) | 0.15 | 0.495 | 1.2575 | 3.04 |
|  | Office worker | 0.1 | 29 | <LOD | <LOD | <LOD | 0.1275 | 0.56 |
| **BDzP** |  |  |  |  |  |  |  |  |
|  | Firefighter | 0.2 | 7 | <LOD | <LOD | <LOD | <LOD | 0.22 |
|  | Office worker | 0.2 | 0 | <LOD | <LOD | <LOD | <LOD | <LOD |
| **DoCP** |  |  |  |  |  |  |  |  |
|  | Firefighter | 0.1 | 9 | <LOD | <LOD | <LOD | <LOD | 0.15 |
|  | Office worker | 0.1 | 1 | <LOD | <LOD | <LOD | <LOD | <LOD |
| **DpCP** |  |  |  |  |  |  |  |  |
|  | Firefighter | 0.1 | 41 | <LOD | <LOD | <LOD | 0.2275 | 0.45 |
|  | Office worker | 0.1 | 17 | <LOD | <LOD | <LOD | <LOD | 0.20 |
| **TBBA** |  |  |  |  |  |  |  |  |
|  | Firefighter | 0.2 | 24 | <LOD | <LOD | <LOD | <LOD | 0.42 |
|  | Office worker | 0.2 | 8 | <LOD | <LOD | <LOD | <LOD | 0.24 |
| **TBBPA** |  |  |  |  |  |  |  |  |
|  | Firefighter | 0.2 | 45 | <LOD | <LOD | <LOD | 0.41 | 0.96 |
|  | Office worker | 0.2 | 42 | <LOD | <LOD | <LOD | 0.29 | 0.64 |
| Abbreviations: LOD = Limit of Detection; DF = Detection Frequency; GM = Geometric mean; GSD = geometric standard deviation  ^a^2,2',4,4'-Tetrabromodiphenyl ether (OH-BDE 47) and 1,3,5-Tribromo-2-(2,4-dibromophenoxy)benzene (OH-BDE 100) were also quantified but not detected in participant’s samples. | | | | | | | | |

| **Table S2.** β(95%CI) and percent change (95% confidence intervals) in TSH levels for each doubling of exposure of continuous BDCPP, BCEP and DBuP in the full cohort in adjusted ^a^ OLS regression models | | | |
| --- | --- | --- | --- |
|  | **Model** | **β(95%CI)** | **Percent change (95%CI)** |
| Full cohort |  |  |  |
|  | BDCPP | -0.006 (-0.110,0.098) | -0.408 (-7.323,7.024) |
|  | BCEP | -0.069 (-0.161,0.024) | -4.657 (-10.572,1.650) |
|  | DBuP | -0.015 (-0.137,0.107) | -1.033 (-9.035,7.673) |
| Firefighters |  |  |  |
|  | BDCPP | 0.050 (-0.150,0.251) | 3.548 (-9.875,18.972) |
|  | BCEP | -0.127 (-0.297,0.043) | -8.418 (-18.610,3.049) |
|  | DBuP | 0.018 (-0.195,0.231) | 1.257 (-12.658,17.388) |
| Office workers ^b^ | |  |  |
|  | BDCPP | -0.022 (-0.141,0.098) | -1.491 (-9.325,7.020) |
| ^a^ Models adjusted for age and log(creatinine) values below LOD replaced with LOD/sqrt(2); ^b^ Among office workers only BDCPP had DF >70% to use in linear models. | | | |

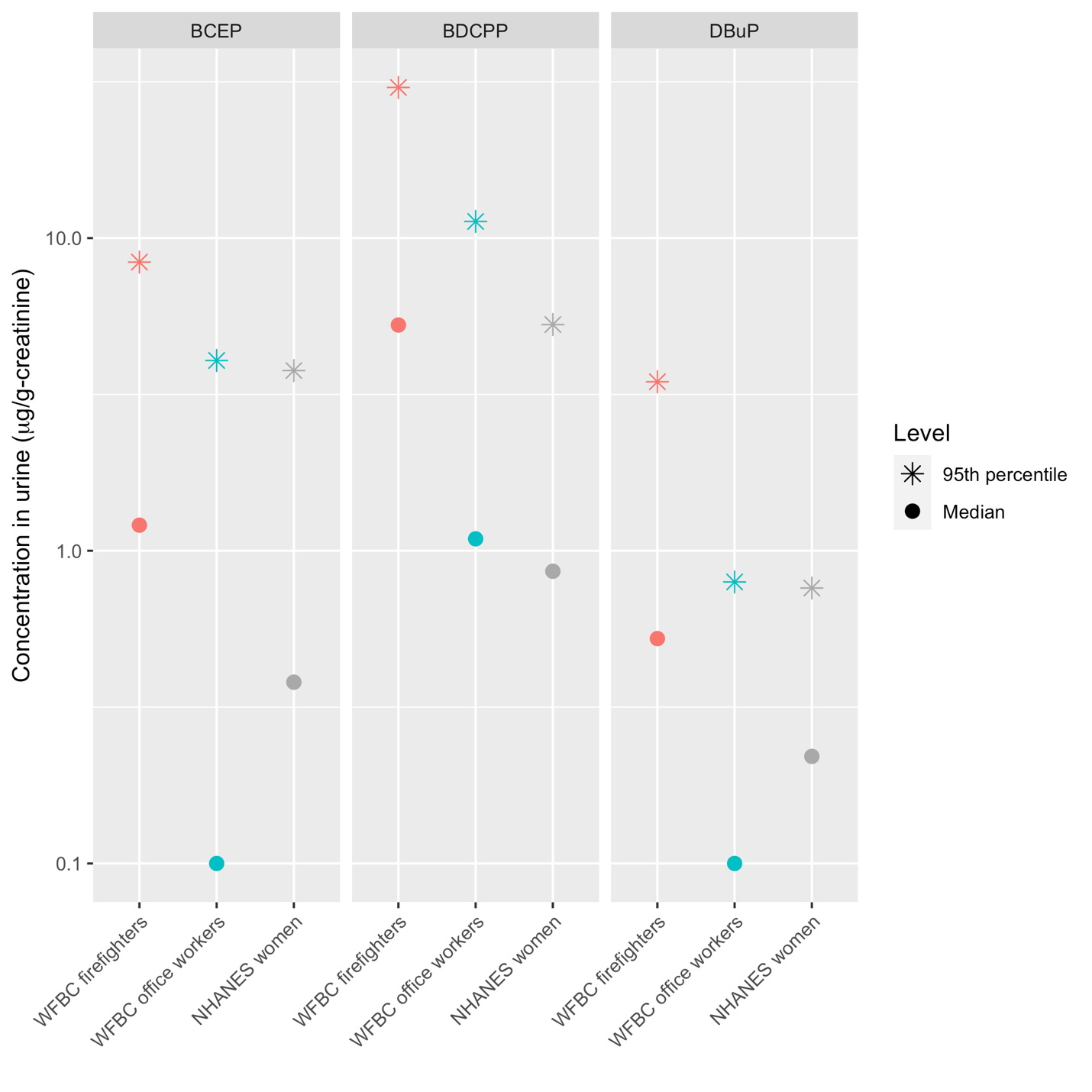

**Figure S1**. Median and 95th percentiles of flame retardants measured in urine (ug/g-creatinine) from 2014-2015 WFBC firefighters and office workers compared to 2013-2014 NHANES adult women (ages 18 to 65) for compounds with at 70% DF among firefighters of the WFBC.
